# Predicting pediatric stroke outcomes using adult lesion-symptom models

**DOI:** 10.64898/2026.11.25.26368989

**Authors:** Joseph C. Griffis, Joel Bruss, Asmaa Mhanna, Michal Brzus, Heather J. Fullerton, Max Wintermark, Nomazulu Dlamini, Nancy K. Hills, Michael M. Dowling, Warren Lo, Timothy J. Bernard, Lori C. Jordan, Mark Mackay, Laura Lehman, Adam Kirton, Dana D. Cummings, Sarah Lee, J. Michael Taylor, Christine K. Fox, Aaron D. Boes, SCOUTS Investigators

## Abstract

Lesion location is a key predictor of post-stroke outcomes in adults, but its relevance for predicting childhood stroke outcomes is unclear. Recent developmental neuroscience work suggests that the gross spatial organization of large-scale brain systems may be generally conserved across development, raising the possibility that damage to common anatomical substrates might underlie chronic domain-level impairments after both adult-onset and childhood stroke. If so, then it should be possible to leverage adult lesion-symptom relationships to predict chronic childhood stroke outcomes. We tested the hypothesis that lesion-symptom models trained to predict sensorimotor and language impairments in patients with adult-onset brain lesions can be translated to predict chronic sensorimotor and language outcomes after childhood stroke.

We trained multivariate lesion-symptom models to predict sensorimotor (Grooved Peg-Board Test: *N*=463; median age=53.6, IQR=17.9 years) and language (Token Test: *N*=379; median age=53.2, IQR=19.4 years) impairments in patients with adult-onset brain lesions. We applied the trained models to lesion segmentations from childhood stroke survivors enrolled in the SCOUTS Study (*N*=469; median age=9.3, IQR=11.0 years), resulting in outcome predictions that we compared against chronic (6+ month) measures of sensorimotor and language impairment derived from the Pediatric Stroke Outcome Measure and the Recurrence and Recovery Questionnaire.

The adult lesion-symptom models generated outcome predictions that robustly correlated with pediatric impairment scores (sensorimotor: rho=0.43; language: rho=0.42; *P*_FWE_ <0.001), enabled classification of pediatric impairment status (both AUC=0.76; *P*_FWE_=0.004), and afforded unique predictive power in multivariable pediatric outcome models (sensorimotor: adjusted OR=3.6, 95% CI=2.4-5.2; language: adjusted OR=3.0, 95% CI=1.67-5.30). Lesion-symptom models trained using pediatric lesion and outcome data captured robust lesion-symptom relationships (both rho_cv_=0.40, *P*<0.001), but yielded no advantage over the adult model predictions when compared directly via cross-validation in the pediatric cohort. Further, the adult and pediatric models made similar predictions when applied to the same lesion data even after adjusting for common effects of lesion size and laterality, suggesting a general convergence towards common anatomical signals present in both children and adults.

Lesion-symptom models trained in patients with adult-onset brain lesions can be translated across outcome measures and developmental stages to predict chronic sensorimotor and language impairments after childhood stroke. These results establish lesion location as an important predictor of childhood stroke outcomes, demonstrate the feasibility of translating adult lesion-symptom models for chronic outcome prediction in children, and support the broader conclusion that the critical anatomy underlying post-stroke sensorimotor and language impairments may be generally conserved across development.

## Introduction

Long-term outcomes vary substantially among children who survive an arterial ischemic stroke^1–4^, and nearly 25% of childhood stroke survivors exhibit moderate-to-severe neurological impairments two years after stroke onset^3^. This variability has been linked to multiple factors including age at stroke onset^3,5,6^, lesion size^7,8^, the affected vascular distributions^6^, and the presence of underlying chronic disease conditions^2,8,9^. However, findings reported in the research literature are mixed and often inconsistent^9,10^, and clinicians remain limited in their ability to predict which children will experience chronic impairments in critical domains such as sensorimotor and language function^9^, creating substantial uncertainty for patients and caregivers and potentially leading to missed opportunities for early intervention.

Lesion location is an important determinant of chronic outcomes after adult-onset stroke^11–15^, and machine learning models trained to capture lesion-symptom relationships in patients with adult-onset brain lesions can predict outcomes in independent samples of adult stroke survivors^13–15^. However, large-scale studies linking lesion location to childhood stroke outcomes are scarce, and the value of lesion location as a predictor of childhood stroke outcomes is unclear^3,6,9,10,16–19^. Given the cost and difficulty of collecting high-quality childhood stroke datasets, it would be of great benefit if adult lesion-symptom models could be directly translated to pediatric applications. While it cannot be assumed that the lesion-symptom relationships identified in adults will generalize to children due to developmental differences in brain organization and response to injury^1,9,10^, there is evidence that the gross spatial organization of large-scale brain networks may largely resemble that observed in adults^20^ by two years of age^21^ or earlier^22,23^, suggesting that the core anatomical infrastructure underlying behaviorally relevant networks may be generally conserved from early childhood to adulthood.

Recent work by our group found that lesion locations associated with post-stroke epilepsy in adults also confer increased risk of seizures and epilepsy in children^24^, demonstrating that at least some lesion-symptom relationships are conserved across development. We also observed that lesion locations associated with chronic sensorimotor and language impairments in childhood stroke survivors appear generally consistent with those documented in adults^25^, supporting the plausibility of translating adult lesion-symptom models to pediatric applications. However, statistically significant lesion-symptom associations are not guaranteed to confer meaningful predictive utility at all^26–30^, much less when generalizing lesion-symptom models across clinical populations and developmental stages. Further, matched outcome measures may often not be available for adult and pediatric patient samples, making cross-outcome model generalization an additional *de facto* pre-requisite for successful and practically useful model translation. While our prior results are encouraging, the feasibility of generalizing this translational approach beyond pathological outcomes such as post-stroke epilepsy remains unclear and must be determined empirically, motivating the current study.

Here, using lesion and chronic outcome data obtained from the Iowa Lesion Registry and the Seizure and Children’s Outcomes after Stroke (SCOUTS) study^24^, we aimed to comprehensively test the hypothesis that adult lesion-symptom models can be translated to predict chronic sensorimotor and language outcomes after childhood stroke. Specifically, we hypothesized that outcome predictions generated by applying adult lesion-symptom models to pediatric lesion data would (1) show robust correlations with chronic pediatric impairment scores, (2) enable classification of chronic pediatric impairment status, and (3) provide unique predictive value even after accounting for other factors such as age at stroke onset, left and right hemispheric lesion volumes, and time from stroke onset to chronic assessment. We also explored whether lesion-symptom models trained in adults vs. children captured common or divergent structure-function relationships.

## Materials and methods

### Adult datasets

Adult datasets were obtained from the Iowa Lesion Registry. Patients were included in the Registry if they had focal acquired brain lesions, neuropsychological testing 3 months or more after the onset of the lesion, and research-quality structural neuroimaging. Patients were excluded from the Registry if they had pre-existing intellectual disability or neurological/psychiatric disorders. Patients with performance data for the Grooved Peg-Board Test (*N*=463) or the Token Test (*N*=379) were included in this study. Patients with surgical resections for epilepsy were excluded. Clinical and demographic characteristics of the adult datasets are summarized in **Table 1**.

**Table 1.** Demographic and clinical characteristics of the adult patient samples.

| **Characteristics** | **Grooved Peg-Board (*N*=463)** | **Token Test (*N*=379)** |
|---|---|---|
| Lesion Onset Age, median (IQR) | 53.6 (17.9) | 53.2 (19.4) |
| Male Gender, *n* (%) | 244 (53) | 205 (54) |
| Race, *n* (%) |  |  |
| White | 450 (97) | 363 (96) |
| Black | 5 (1) | 4 (1) |
| Other | 8 (2) | 12 (3) |
| **Lesion Etiology, *n* (%)** |  |  |
| Ischemic Stroke | 252 (54) | 225 (60) |
| Hemorrhagic Stroke | 88 (19) | 77 (20) |
| Tumor Resection | 55 (12) | 42 (11) |
| AVM Resection | 17 (4) | 19 (5) |
| Other | 51 (11) | 16 (4) |
| **Lesion Laterality, *n* (%)** |  |  |
| Left | 193 (42) | 208 (55) |
| Right | 173 (37) | 106 (28) |
| Bilateral | 97 (21) | 65 (17) |
| **Lesion volume (mean ± SD)** |  |  |
| Total | 35171±51763 | 45334±57008 |
| Left | 16510±35806 | 25248±40796 |
| Right | 18647±38463 | 20065±46816 |

Note – Lesion volume estimates correspond to 1mm<sup>3</sup> resolution voxel counts in template space. Hemispheric lesion volume estimates shown in the table were computed only for patients with damage to the relevant hemisphere.

### Pediatric dataset

The pediatric dataset was obtained from a large multi-site pediatric cohort of 528 children (age 29 days to 18 years) with confirmed arterial ischemic stroke who were prospectively enrolled in either the VIPS^31^ or VIPS II^32^ studies. Chronic stroke outcomes were collected in the Seizures and Children’s Outcomes after Stroke (SCOUTS) study^24^. Clinical imaging was collected and arterial ischemic stroke was centrally confirmed via the VIPS studies; long-term stroke outcomes were collected through the SCOUTS study. Neonates were not included in the parent studies (VIPS I and II). Children with chronic (>=6-month post-onset) neurological assessment data and lesion segmentations obtained from neuroimaging scans were included here (*N*=469). Clinical and demographic characteristics of the pediatric dataset are summarized in **Table 2**.

**Table 2.** Demographic and clinical characteristics of the pediatric stroke sample.

| **Characteristics** | **Pediatric Stroke (*N*=469)** |
|---|---|
| Stroke Onset Age median (IQR) | 9.3 (11.02) |
| Years to Chronic Assessment, median (IQR) | 3.32 (4.26) |
| Male, *n* (%) | 266 (57) |
| Race, *n* (%) |  |
| White | 320 (68) |
| Black | 58 (12) |
| Asian | 22 (5) |
| Other | 69 (15) |
| **Stroke Classifications, *n* (%)** |  |
| Idiopathic | 25 (5) |
| Cardioembolic | 18 (4) |
| Definite Arteriopathy | 177 (38) |
| Possible Arteriopathy | 92 (20) |
| No Arteriopathy | 157 (33) |
| **Comorbidities, *n* (%)** |  |
| Chronic Heart Disease | 69 (15) |
| Endocarditis | 5 (1) |
| Head Trauma | 45 (10) |
| Meningitis | 21 (5) |
| Sickle Cell | 16 (3) |
| Systemic Sepsis | 18 (4) |
| **Lesion Laterality, *n* (%)** |  |
| Left | 180 (38) |
| Right | 193 (41) |
| Bilateral | 96 (21) |
| **Lesion Volume (mean ± SD)** |  |
| Total | 63999±107245 |
| Left Hemisphere | 52939±85387 |
| Right Hemisphere | 53304±100620 |

Note: Lesion volume estimates are computed as counts using 1mm isotropic voxels. Means and standard deviations for left and right hemispheric lesion volumes were computed only for patients with damage to each hemisphere.

### Standard protocol approvals, registrations, and patient consents

The SCOUTS study was approved by an Institutional Review Board (IRB) at Vanderbilt University Medical Center, which served as the IRB of record for all participating domestic sites. Local IRB approved the study at all participating sites outside the United States in the SCOUTS study and at each enrolling site for the VIPS and VIPSII studies. This analysis was approved by the University of Iowa Institutional Review Board (Project ID 201705278). Written informed consent was obtained from all participants or parents of children, as applicable, in accordance with the Declaration of Helsinki. In some instances, chart review was performed for follow-up data in participants that had previously consented to research, and the need for re-consenting participants was waived as the research was deemed minimal risk. Follow-up data were collected through chart review with a waiver of consent for minimal risk research, or through standardized parental questionnaires.

### Cognitive and behavioral measures

#### Adult outcomes

The Grooved Peg-Board Test was used to define a measure of sensorimotor impairment by subtracting the *t*-score for the body side contralateral to the lesion from the *t*-score for the body side ipsilateral to the lesion, such that larger difference scores reflect worse performance for the contralateral body side. For patients with bilateral lesions, the smaller of the two *t*-scores was subtracted from the larger *t-*score. Difference scores were used to reduce the influence of inter-individual performance variability while amplifying variability attributable to the lesion, providing a single impairment score that could be jointly analyzed in all patients regardless of lesion laterality. The Grooved Peg-Board Test was chosen because it is the only sensorimotor measure available for many patients in the Iowa Registry, and because it correlates with chronic sensorimotor impairment scores from the Pediatric Stroke Outcome Measure (PSOM)^33^, which was used to define the pediatric outcomes here.

The Token Test was used as a measure of language impairment. It is a sensitive measure of auditory language comprehension commonly used to screen for aphasic disorders. As noted below, expressive and receptive language scores were highly correlated in the pediatric dataset, leading us to collapse them into a single measure of general language impairment. Therefore, we used a single adult language outcome for the primary analyses. The Token Test was selected because it is well-represented among our adult patient sample, it has been shown to be highly effective for identifying aphasic patients regardless of clinical sub-type^34^, and because multivariate lesion-symptom models of Token Test performance have been previously found to implicate distributed left fronto-temporal structures consistent with the broader language system^13,14,35^.

#### Pediatric outcomes

Chronic sensorimotor and language outcomes were assessed using the PSOM^33^. The PSOM is a validated measure of neurological impairment based on clinical assessment that scores deficits in five domains: right and left sensorimotor, expressive language, receptive language and cognitive/behavioral^33^. When PSOM data were not available, the related Recurrence and Recovery Questionnaire (RRQ)^36^, a parent-administered assessment that correlates strongly with the PSOM^36^, was used. Both assessments are scored on a 4-point ordinal scale. Higher scores indicate greater impairment.

We defined two primary outcomes corresponding to measures of chronic sensorimotor and language function. Mirroring the adult sensorimotor outcome, the pediatric sensorimotor outcome was defined as the sensorimotor score corresponding to the body side contralateral to the lesion; for bilateral lesions, the worst of the two scores was used. The expressive and receptive language sub-domain scores were highly correlated (Spearman rho=0.73, *P*<0.001), leading us to define a single summary outcome as the worst score across the two sub-domains. This allowed us to retain the 4-point score structure of the original measures, while summing or averaging the scores would not, preserving consistency with the PSOM/RRQ, which are commonly used for both clinical and research purposes, and with the sensorimotor outcome, allowing identical analysis strategies for each outcome. The “worst” language measure strongly correlated with the sum of the sub-domain scores (Spearman rho=0.99, *P*<0.001), consistent with a measure of general language impairment.

### Neuroimaging data

Detailed descriptions of the neuroimaging data collection and image processing workflows for the adult^13,14^ and pediatric^24^ cohorts are reported elsewhere.

For the pediatric cohort, clinical imaging and clinical presentation data were independently reviewed by a board-certified neuroradiologist (M.W.) and a pediatric vascular neurologist (H.J.F.) to confirm stroke. Typical pediatric stroke MRI protocols included T1, FLAIR, gradient echo, diffusion weighted imaging (DWI), and apparent diffusion coefficient (ADC) sequences. Lesions were manually traced in native subject space using FSL, with primary reliance on DWI and ADC. The native MRI and co-registered lesion mask were linearly aligned and non-linearly warped to an MNI152 template brain using ANTs. Enantiomorphic normalization^37^ was used to improve registration for unilateral lesions, and cost-function masking^38^ was used for bilateral lesions. Lesion segmentation and template registration quality was reviewed by experts, including A.D.B. and J.B., and any errors were manually corrected. Investigators involved in lesion tracing and editing were blind to the outcomes. Total and hemispheric lesion volumes were estimated as 1mm isotropic resolution voxel counts in template space since this removes variability due to factors such as head size or brain volume. Lesion segmentations were resampled to 2mm isotropic voxel resolution prior to lesion-symptom modeling analyses.

### Statistical analyses

Statistical analyses were conducted using MATLAB R2025b (The MathWorks).

#### Multivariate models – adult datasets

We trained separate partial least squares regression (PLSR) models to predict Grooved Peg-Board and Token Test outcomes based on lesion location using the Iowa Brain-Behavior Modeling Toolkit^35^, available at (https://github.com/jcgriffis/ibb_toolkit), similar to prior work^14,35,39^. Five repetitions of nested 5-fold cross-validation were used for initial model evaluation, with the inner loop used to optimize the number of PLS components, and the outer loop used to evaluate prediction performance^14,35^. Cross-validation folds were stratified according to the quartiles of the outcomes to ensure adequate representation of impairment levels across training and test folds^14,35^. Cross-validation performance was summarized by computing the full-sample cross-validation correlation between the average (across repetitions) test fold predictions and the true observed scores^35,40^. A final model was trained on the full adult dataset for each outcome using five repetitions of 5-fold cross-validation to optimize the number of PLS components, and the fully-trained model was applied to the pediatric lesion data to obtain out-of-sample predictions for each child. For the Grooved Peg-Board model, all lesions were rendered symmetric by flipping across the midline to increase power, analogous to other work^25,41–43^. For patients with bilateral lesions, the flipped lesions included voxels that were lesioned in either hemisphere.

For our primary analyses, we generated predictions using only positive model weights (i.e. voxels predicting impairment), motivated by the consideration that the “symptom-positive” anatomy aligns with the expected direction of lesion-symptom causality, and by the concern that “symptom-negative” model weights for lesion sites associated with good outcomes in the training sample may reflect overfitting driven by systematic spatial biases in the lesion locations associated with good outcomes^44^. That is, since the existence of “critical regions” where damage causes good outcomes is usually implausible^44,45^, we focused on only voxels where damage predicted deficits. For completeness, results obtained using the full model weights are reported in the supplement.

#### Multivariate models – pediatric dataset

We trained and evaluated pediatric PLSR lesion-symptom models analogously to the adult models. Since this required us to model the ordinal outcomes as continuous variables, we re-coded them to have equal intervals between each level. We used a regression, rather than classification, approach since this provided consistency between the adult and pediatric models and allowed the models to learn more granular lesion-symptom relationships by preserving the full range of impairment severities during training. Since only deficit-predicting voxels were used to generate the adult model predictions in the pediatric dataset, we mirrored this during cross-validation of the pediatric models to ensure that the test fold predictions could be directly compared against the adult model predictions; test fold predictions generated using only the deficit-predicting model weights were re-scaled to the appropriate score range using an intermediate linear regression model fit within the training folds. Analyses using full model weights are also reported in the supplement. As with the adult models, full-sample cross-validation correlations were computed to summarize cross-validation performance, but we report them as both linear and rank correlations due to the ordinal outcome scales. As with the adult Grooved Peg-board model, lesions were rendered symmetric prior to training the contralateral sensorimotor outcome model.

#### Hypothesis-driven analyses

First, we tested whether the pediatric outcome predictions made by the adult lesion-symptom models correlated with the true pediatric outcome scores by computing Spearman rank correlations between the predicted and observed scores. We also computed partial rank correlations while covarying left and right hemispheric lesion volumes, age at stroke onset, and time from onset to neurological assessment to verify that associations could not be fully attributed to other variables. Statistical significance was evaluated using parametric *P*-values, and bootstrapping (1000 iterations) was used to compute 95% confidence intervals (CIs).

Second, we tested whether the predictions made by the adult lesion-symptom models could be used to classify pediatric impairment status as “unimpaired” (i.e. scores of 0 or 0.5) vs. “impaired” (i.e. scores of 1 or 2). We constructed receiver-operating characteristic (ROC) curves using the adult model predictions and impairment labels, and computed the area under the curve (AUC)^28^. These analyses were repeated after regressing the same covariates described above out of the adult model predictions. Statistical significance was evaluated via permutation testing (1000 iterations), and bootstrapping (1000 iterations) was used to compute 95% CIs.

Third, we fit multivariable ordinal logistic regression models to estimate cumulative adjusted odds ratios (aOR) associated with the severity of impairments predicted by the adult models while accounting for the effects of other factors. This involved first fitting “base” ordinal logistic regression models in the pediatric sample, with the covariates described above as the initial predictor set. Then, we used likelihood ratio tests to evaluate the change in model fit resulting from adding the adult model predictions to these “base” models, and we estimated aORs along with 95% CIs for each of the standardized predictors in the models.

Since the above analyses correspond to different tests of the same research hypothesis, i.e. that adult lesion-symptom models can predict chronic childhood stroke outcomes, Bonferroni-Holm correction was used to control the family-wise error rate at 0.05 across all of the above tests.

Since we previously identified a moderating effect of age at stroke onset on the relationship between left hemispheric lesion volumes and expressive language impairments^25^, we verified that results for the language outcome did not differ when the “base” model included this moderation effect. We also explored whether age uniquely moderated the association between impairment severity predicted by the adult model and chronic language outcomes by performing an additional analysis in which an interaction term between the adult model predictions and age was added to the “base” model that already included the moderation effect for left hemisphere lesion volume, and we tested whether this improved the model fit using a likelihood ratio test.

Finally, we used a cross-validation approach to test whether training lesion-symptom models directly within the pediatric dataset conferred an advantage over simply applying the adult-trained models to the pediatric lesion data. Logistic ridge regression models classifying pediatric impairment status were trained and evaluated using an analogous cross-validation procedure as for the lesion-symptom models. Here, we used classification models, rather than regression models, since classification performance is better-aligned with how lesion-informed prognostication models would likely be applied in clinical settings^14^, but results obtained using regression approaches are reported in the supplement. For each outcome, we trained and evaluated a “base” model that included the same predictors as the “base” ordinal logistic regression models described above along with “expanded” models that additionally included either (a) the domain-matched adult lesion-symptom model predictions or (b) the test fold predictions obtained from cross-validation of the domain-matched pediatric lesion-symptom models (matched by fold and repetition). Cross-validation partitions were identical for all models, including the lesion-symptom models, to allow direct comparison of performance estimates. To reduce the impact of class imbalances, misclassification costs for impaired patients were set to be proportional to the magnitude of the class imbalance as in prior work^14,35^. Dependent samples *t*-tests were used to compare average test fold AUCs between each pair of models^14,35^. Bonferroni-Holm correction was applied to the results of the cross-validation analyses.

#### Post-hoc analyses

We computed partial Pearson correlations (i.e. covarying hemispheric lesion volumes) between (a) the predictions obtained by applying the adult lesion-symptom models to the pediatric lesion data and (b) the test fold predictions obtained from cross-validation of the pediatric lesion-symptom models to evaluate the similarity of model predictions generated using the same lesion inputs, and we tested whether they were more similar for domain-matched vs. domain-mismatched models using Steiger *z*-tests. We also computed spatial correlations to evaluate the similarity of hemispheric weight topographies encoded by the adult and pediatric lesion-symptom models for the language outcome. These analyses were considered descriptive, and so they are reported with uncorrected *P*-values.

## Results

### Lesion and outcome data

Voxel-wise lesion frequencies for each adult brain lesion dataset are shown in **Figure 1A**, and for the pediatric stroke dataset in **Figure 1B**, demonstrating extensive lesion coverage for all datasets.

**Figure 1.**
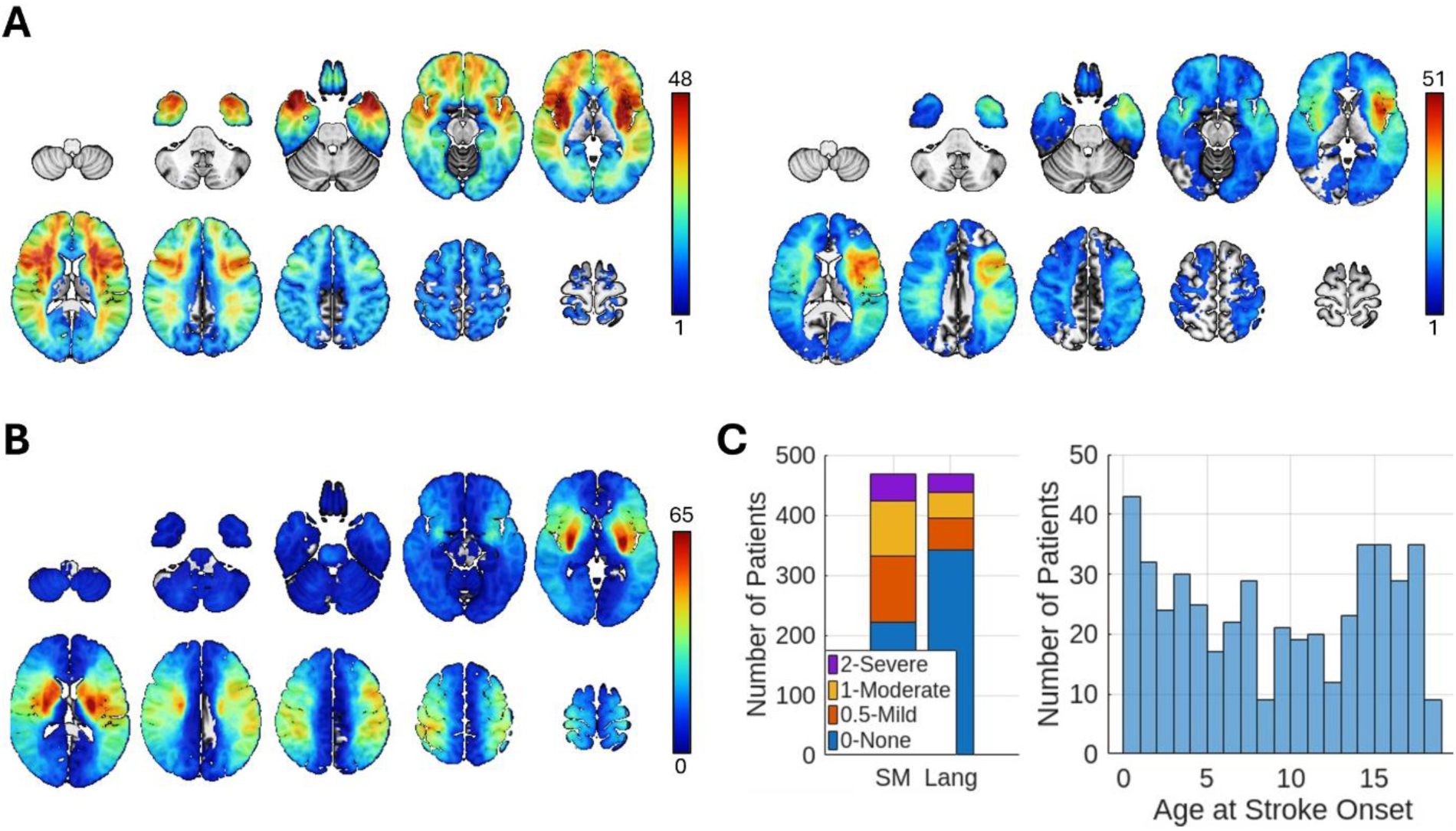
Lesion and behavioral data. **A.** Lesion overlap map for the adult Grooved Peg-board dataset (left, *N*=463) and the adult Token Test dataset (right, *N*=379). Note that the adult dataset used for the Grooved Peg-Board analysis featured lesions that were rendered symmetrical across the midline, causing the overlap map to be mirror symmetric. **B.** Lesion overlap map for the pediatric stroke dataset (*N*=469). Note that flipped lesions were used when generating pediatric outcome predictions from the adult Grooved Peg-Board model and when training and evaluating the pediatric sensorimotor model (not shown). **C.** PSOM score distributions for the sensorimotor (SM) and language (Lang) outcomes in the pediatric stroke dataset are shown in the stacked bar plot (left), and stroke onset age distributions in the pediatric stroke dataset are shown in the histogram (right). Slices are shown in radiological convention (i.e. left is on the right).

Roughly half (53%) of the pediatric patients exhibited at least mild impairment (i.e. PSOM > 0) on the sensorimotor outcome, while nearly one-third (27%) of the pediatric patients exhibited at least mild impairment (i.e. PSOM > 0) on the language outcome (**Figure 1C**). Ages at stroke onset varied widely in the pediatric stroke dataset, effectively sampling across the full childhood stroke age range (i.e. 29 days to 18 years of age; **Figure 1C**). Outcome assessments were mostly obtained within the first 4 years of stroke onset^25^, and 98% (*n*=459) of children were 2 years or older at the time of assessment.

### Cross-validation of adult lesion-symptom models

We used cross-validation to evaluate the adult lesion-symptom model performance in the adult datasets. The correlations between the predicted and observed outcomes were statistically significant for both adult lesion-symptom models with moderate-to-large effect sizes (Grooved Peg-Board: *r*_cv_*=*0.36, *P*<0.001; Token Test: *r_cv_=*0.71, *P*<0.001), indicating that the models captured robust lesion-symptom relationships that successfully generalized to held-out patients during cross-validation. Summaries of the full weight model topographies are shown in **Figure 2**, emphasizing structures canonically associated with sensorimotor and language function, respectively.

**Figure 2.**
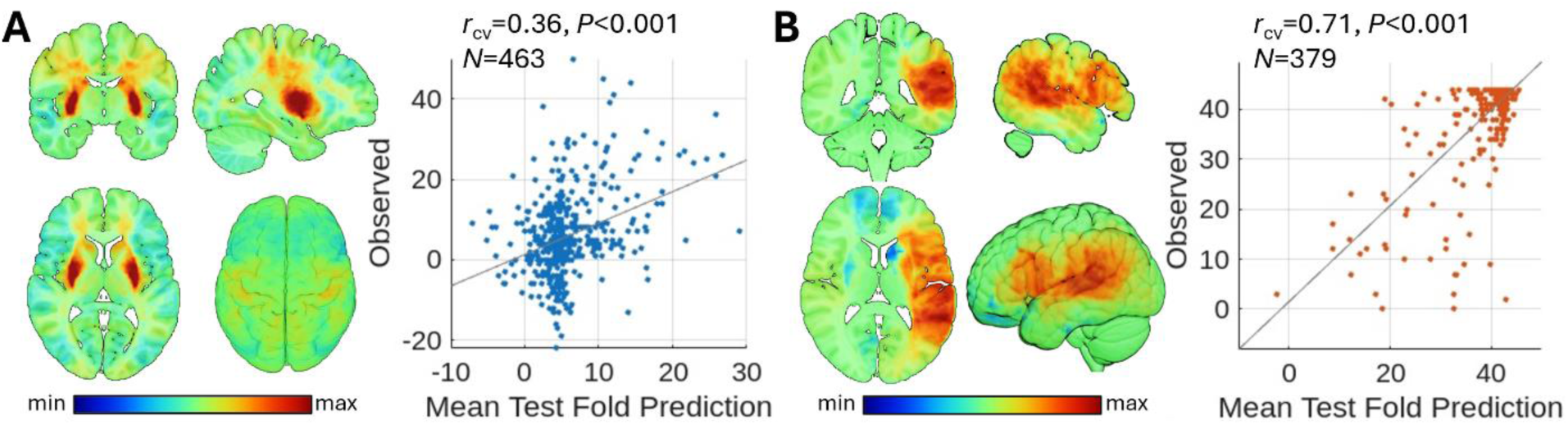
Adult lesion-symptom models. **A-B.** The brain images on the left of each panel summarize the topographies of model weights from the adult lesion-symptom models of sensorimotor (i.e. Grooved Peg-Board Test - **A**) and language (i.e. Token Test - **B**) outcomes. The scatterplots on the right of each panel show the relationships between the average (i.e. across repetitions of the cross-validation procedure) test fold predictions obtained during cross-validation (x-axis) and the true observed scores (y-axis) for each outcome in the adult datasets.The full-sample cross-validation correlations (*r_cv_*) are shown on the top of the plots. Note – while Token Test scores were inverted during model training so that deficit-predicting voxels would have positive signs, the predicted and observed scores were restored to the original score range to visualize the relationship shown in the scatterplot. Slices are shown in radiological convention (i.e. left is on the right).

### Adult-to-pediatric model translation

We applied the deficit-predicting weights from the adult lesion-symptom models (**Figure 3A**) to the pediatric lesion data to generate predicted outcomes for each child. Results obtained using full model weights are reported in **Supplementary Figure 1**.

**Figure 3.**
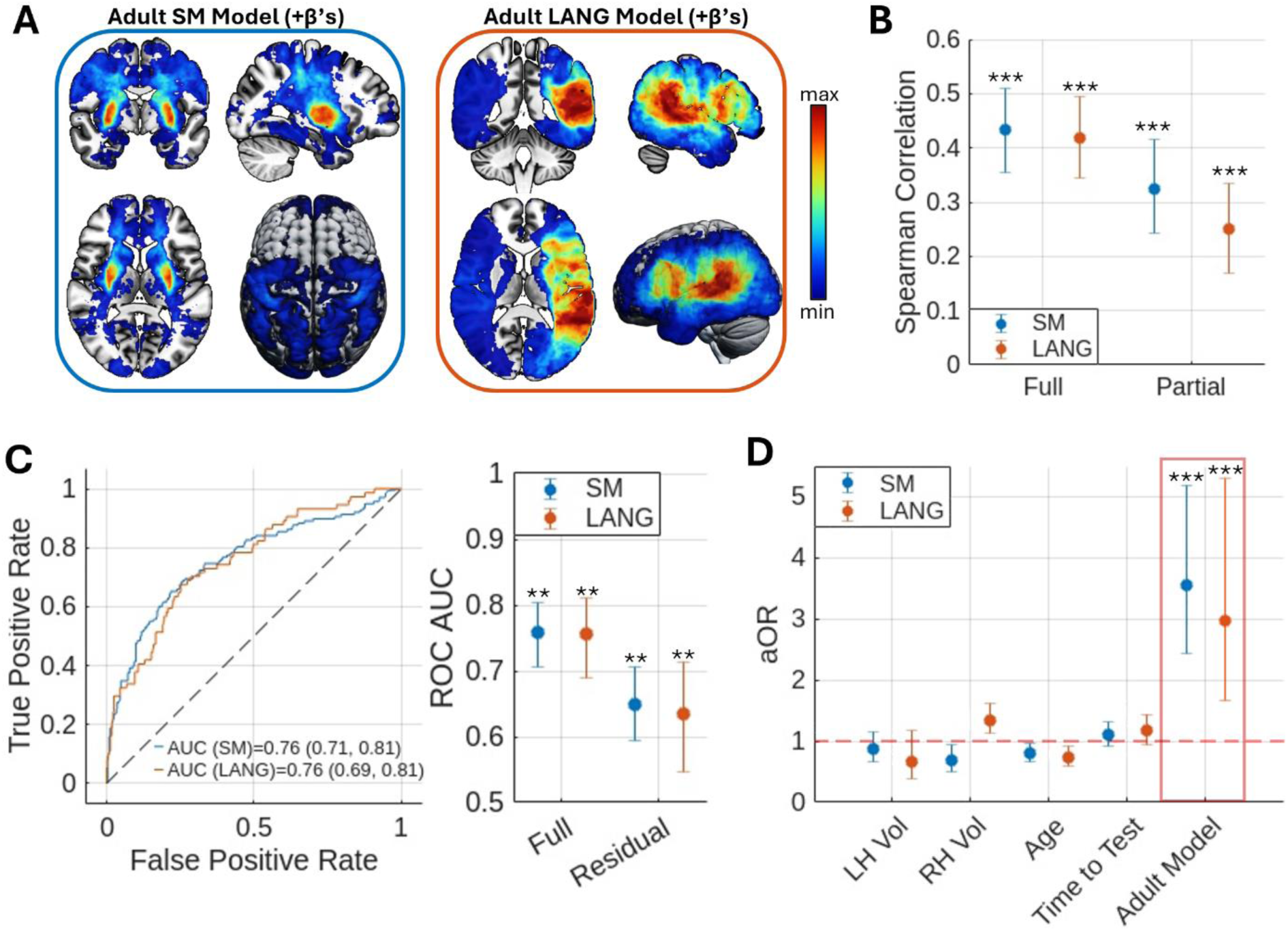
Translating adult lesion-symptom models to predict chronic childhood stroke outcomes. **A**. Brain images show model weights associated with chronic sensorimotor (SM – Grooved Peg-Board Test) and language (LANG – Token Test) impairments in the adult datasets, which were used to generate outcome predictions for the pediatric dataset. Slices are shown in radiological convention (i.e. left is on the right). **B**. Full and partial Spearman rank correlations were computed between the adult model predictions and the true observed sensorimotor (blue – SM) and language (orange – LANG) outcomes in the pediatric dataset. Correlations are shown along with corresponding 95% confidence intervals (CIs). **C.** Left - Receiver-operating characteristic (ROC) curves discriminating between patients with vs. without sensorimotor (blue) and language (orange) impairments were constructed using the adult model predictions. AUCs and 95% confidence intervals (CIs) are shown in the legend. Right – AUCs and 95% CIs are shown for analyses using adult model predictions before (full –same as left panel) and after (residual) regressing out covariates. **D**. Multivariable ordinal logistic regression models were fit to the sensorimotor (blue) and language (orange) outcomes in the pediatric dataset, and the adult model predictions were added as predictor variables (i.e. “expanded” models). Adjusted cumulative odds ratios (aOR) are shown for standardized predictors from the “expanded” models of each outcome (blue – SM, orange – LANG) that included the adult model predictions as predictor variables (i.e. effects of *a priori* interest – red box). \*\*\**P*_FWE_≤0.001, \*\**P*_FWE_≤0.005

The adult model predictions for both the sensorimotor (rho=0.43, 95% CI=[0.36, 0.52], *P*_FWE_<0.001) and language (rho=0.42, 95% CI=[0.35, 0.49], *P*_FWE_<0.001) outcomes were significantly correlated with the observed outcomes (**Figure 3B**), even after adjusting for covariates (sensorimotor: partial rho=0.33, 95% CI=[0.24, 0.41], *P*_FWE_<0.001; language: partial rho=0.25, 95% CI=[0.17, 0.33], *P*_FWE_<0.001), and they enabled statistically significant classification of impairment status for both the sensorimotor (AUC=0.76, 95% CI=[0.71, 0.81], permutation *P*_FWE_*=*0.004) and language (AUC=0.76, 95% CI=[0.69, 0.81], permutation *P_FWE_=*0.004) outcomes (**Figure 3C**) that remained statistically significant when using the covariate-residualized adult model predictions for both the sensorimotor (AUC=0.65, 95% CI=[0.59, 0.71], permutation *P*_FWE_*=*0.004) and language (AUC=0.63, 95% CI=[0.55, 0.71], permutation *P*_FWE_*=*0.004) outcomes.

The addition of the adult model predictions to the “base” ordinal logistic regression models significantly improved model fits for both the sensorimotor (χ^2^[1]=43.70, *P*_FW*E*_<0.001) and language (χ^2^[1]=14.14, *P*_FWE_=0.001) outcomes, with the adult model predictions being identified as statistically significant (*P*_FW*E*_ <= 0.001) predictors in each model (**Figure 3D**, **Supplementary Table 1**). These effects were unique to the domain-matched adult model predictions (**Supplementary Figure 2**), consistent with domain-specific structure-function relationships. Each standard deviation increase in impairment severity predicted by the adult model was associated with ∼3-3.5x greater odds of worse impairment when holding other predictors constant (**Figure 3D**; full model details are reported in **Supplementary Table 1**).

Similar results were obtained when analyses were performed separately for early vs. late childhood stroke survivors (**Supplementary Figure 3**), indicating that our results are applicable to both stroke onset epochs. Similar results were also obtained when the “base” model for the language outcome included an interaction term between age at stroke onset and left hemispheric lesion volume (**Supplementary Figure 4A**). Adding an additional interaction between age at stroke onset and the adult model predictions did not significantly improve the model fit (χ^2^[1]=0.44, *P*=0.51), indicating that the relationship between adult lesion-symptom model predictions and chronic language outcomes did not depend on age at stroke onset when accounting for hemisphere-level interaction effects.

### Comparison of adult vs. pediatric lesion-symptom models

We trained lesion-symptom models in the pediatric cohort. The full-sample cross-validation correlations for the pediatric lesion-symptom models of the sensorimotor (*r_cv_*=0.42, *P*<0.001; *rho_cv_*=0.40, *P*<0.001) and language (*r_cv_*=0.41*, P*<0.001; *rho_cv_*=0.40, *P*<0.001) outcomes were statistically significant with moderate effect sizes, indicating that they exhibited good generalization to held-out patients during cross-validation.

The test fold predictions obtained from cross-validation of the pediatric lesion-symptom models significantly improved cross-validation performance over the “base” models for both the sensorimotor (t[24]=3.42, *P*_FWE_=0.01, 95% CI=[0.01, 0.03]) and language (t[24]=6.34, *P*_FWE_<0.001, 95% CI=[0.02, 0.05]) outcomes, as did the adult model predictions for both the sensorimotor (t[24]=7.81, *P*_FWE_<0.001, 95% CI=[0.04, 0.07]) and language (t[24]=6.99, *P*_FWE_<0.001, 95% CI=[0.03, 0.05]) outcomes (**Figure 4**). However, the improvements afforded by the pediatric lesion-symptom model predictions were significantly less than those afforded by the addition of the adult lesion-symptom model predictions for the sensorimotor (t[24]=-6.73, *P*_FWE_<0.001, 95% CI=[−0.04, −0.03]) outcome (**Figure 4A**), and they were also numerically less for the language outcome (**Figure 4B**) but this difference was not statistically significant (t[24]=-1.09, *P*_FWE_=0.29, 95% CI=[-0.006, 0.002]).

**Figure 4.**
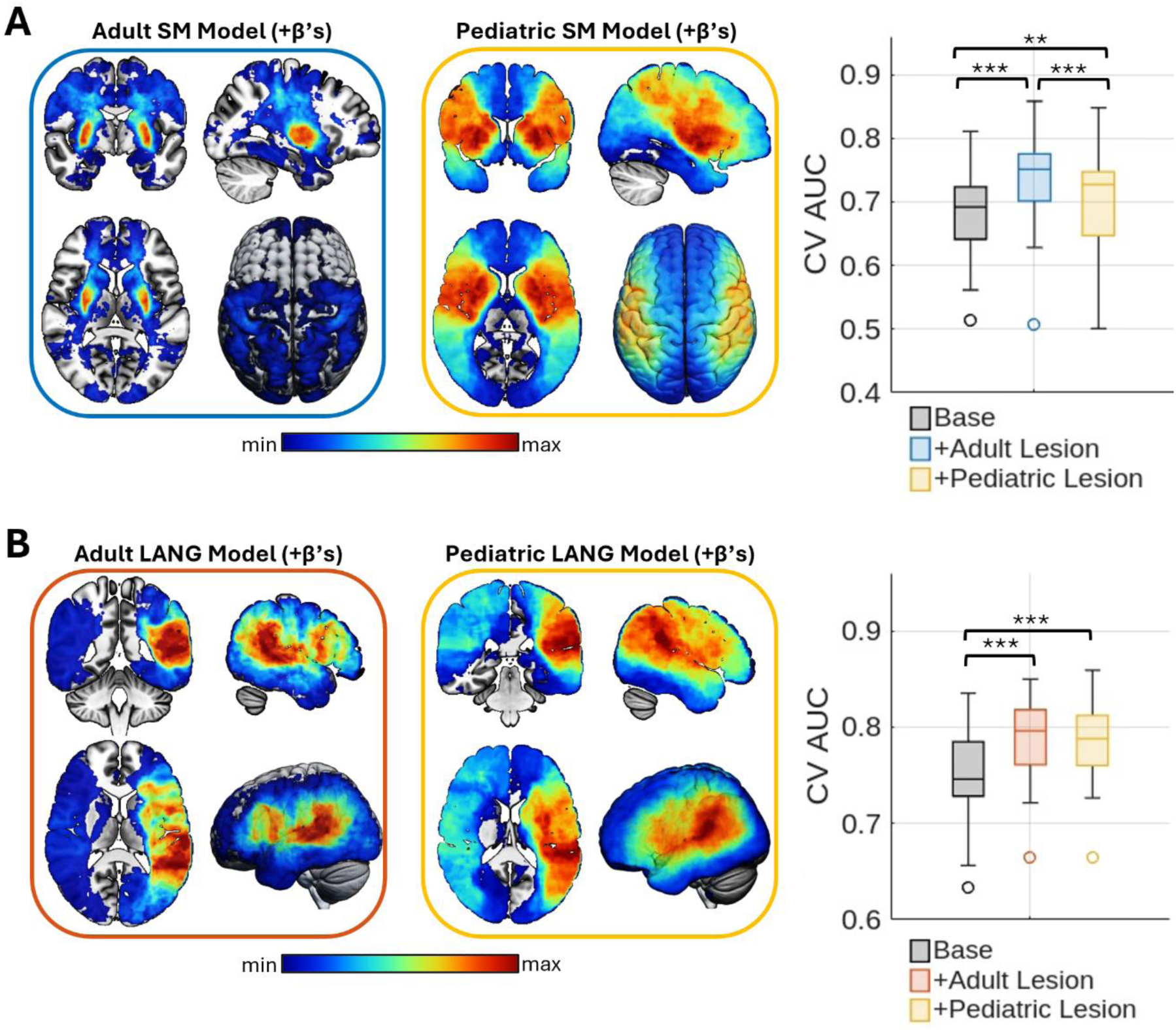
Comparing multivariable models incorporating adult vs. pediatric lesion-symptom model predictions via cross-validation. Cross-validation model comparison results are shown for the sensorimotor (**A**) and language (**B**) outcomes. Brain images show deficit-predicting model coefficients from the final adult (left, blue and orange outlines) and pediatric lesion-symptom models (right, yellow outlines). Box plots show comparisons of cross-validation test fold classification performance (AUC) between the “base” logistic ridge regression models (grey), the “expanded” models that included the adult lesion-symptom model predictions (blue – SM, orange – LANG), and the alternate “expanded” models that instead included the pediatric lesion-symptom model predictions (yellow). Higher AUC values indicate better test fold classification performance. Slices are shown in radiological convention (i.e. left is on the right). \*\*\**P*_FWE_<0.001,\*\**P*_FWE_=0.01

Highly similar results were obtained when using the full model weights to generate predictions (**Supplementary Figure 1E**), when the “base” model for the language outcome included an interaction term between left hemispheric lesion volume and age at stroke onset (**Supplementary Figure 4B**), and when a regression (rather than classification) approach was used (**Supplementary Figure 5**).

### Comparison of model predictions and weight topographies

We compared predictions generated by applying the adult and pediatric models to the pediatric lesion data. Since it is possible that similar model predictions could reflect common but regionally non-specific relationships captured by the adult and pediatric models, we performed these analyses while adjusting for hemispheric lesion volumes (**Figure 5**). The test fold predictions generated during cross-validation of the pediatric lesion-symptom models were highly correlated with the out-of-sample predictions obtained by applying the adult lesion-symptom models to the pediatric lesion data for both the sensorimotor (partial *r*=0.76, *P*<0.001) and language (partial *r*=0.83, *P*<0.001) outcomes. Pediatric model predictions were more strongly correlated with the predictions generated by the domain-matched vs. domain-mismatched adult models (sensorimotor: *z*=3.87, *P*<0.001; language: *z*=11.49, *P*<0.001).

**Figure 5.**
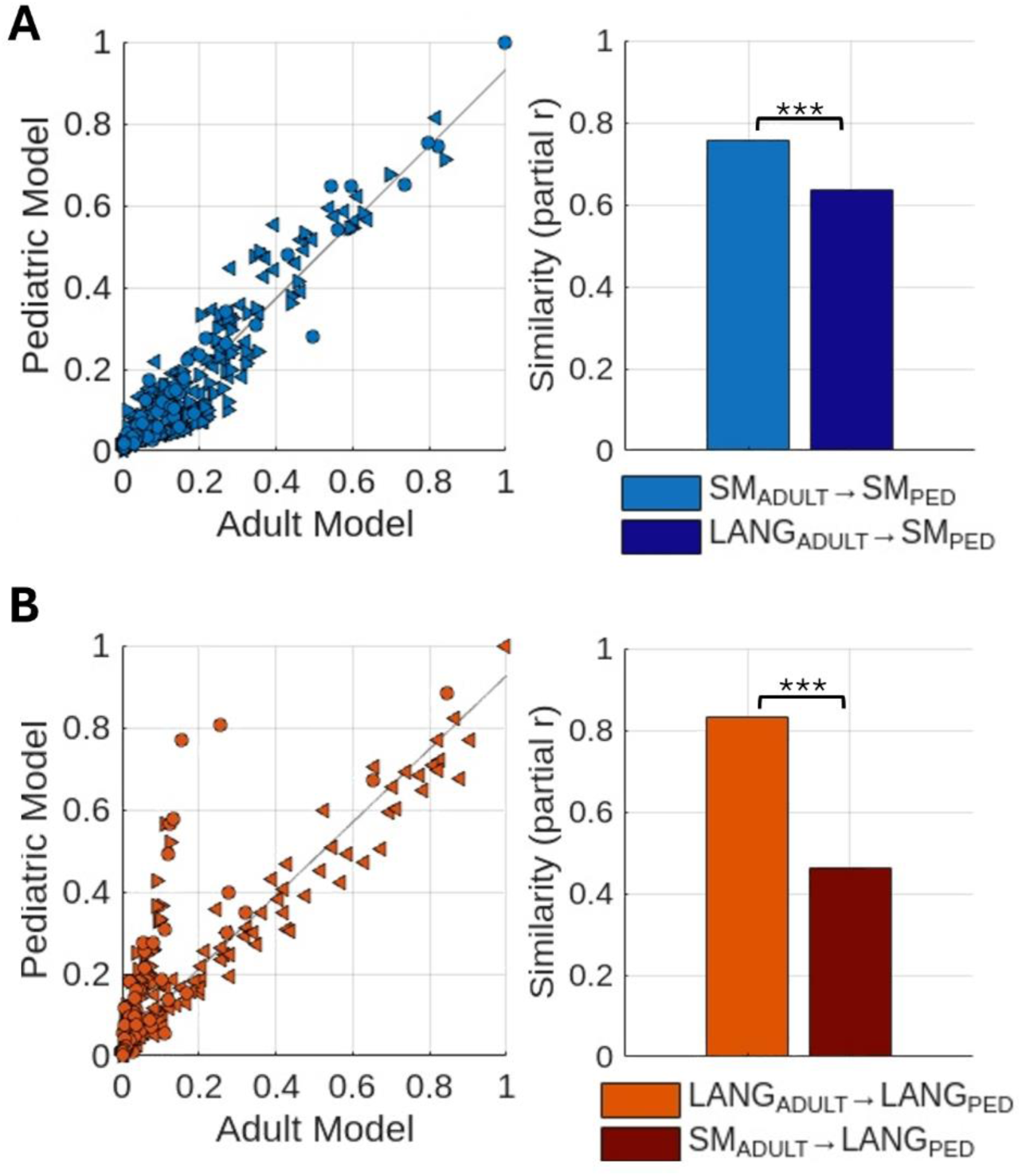
Agreement of outcome predictions generated by applying the adult vs. pediatric lesion-symptom models to the pediatric lesion data. Scatterplots show the relationships between adult lesion-symptom model predictions (x-axes) and pediatric lesion-symptom model predictions (y-axes) of sensorimotor (top - blue) and language (bottom - orange) outcomes. Adult model predictions were obtained by applying the trained adult lesion-symptom models to the pediatric lesion data, while pediatric model predictions were obtained via cross-validation in the pediatric dataset. Each data point represents a single pediatric patient. Patients with left hemispheric lesions are shown with a left-facing arrow (<), patients with right hemispheric lesions are shown with a right-facing arrow (>), and patients with bilateral lesions are shown with a circle. Predictions from each model were rescaled to have the same range [0,1] to allow comparisons of relative predicted impairment severities. Bar plots show partial (i.e. removing shared prediction variance attributable to hemispheric lesion volumes) correlations between adult and pediatric lesion-symptom model predictions obtained using domain-matched (light colored bars) vs. domain-mismatched models (dark colored bars). **A.** Sensorimotor outcome. **B.** Language outcome. \*\*\**P*<0.001

For the language outcome, we were interested in whether the adult and pediatric models made similar predictions regardless of lesioned hemisphere, so we repeated these analyses separately for patients with left vs. right unilateral lesions while adjusting for affected hemisphere lesion volumes. Predictions were highly similar for patients with left unilateral lesions (partial *r*=0.85, *P*<0.001) but less similar for patients with right unilateral lesions (partial *r*=0.46, *P*<0.001), although both were still more similar for the domain-matched vs. domain-mismatched models (left: *z=*8.37, *P*<0.001; right: *z*=4.64, *P*<0.001). Notably, spatial correlations computed between model weights for voxels that were common to both language outcome models also indicated that the left hemispheric model weight topographies of the adult and pediatric models were highly similar (spatial *r*_LH_=0.72) while the right hemispheric model weight topographies were less similar (spatial *r_RH_=*0.22), consistent with the divergence in model predictions for lesions in the right hemisphere.

## Discussion

Major progress has been made towards lesion-informed outcome prediction for adult-onset stroke^12–15^, but much less progress has been made towards leveraging lesion location for predicting outcomes after childhood stroke^1^, likely in part due to the difficulty and cost of collecting large childhood stroke datasets with imaging data suitable for training and testing lesion-symptom models^7^. Our results show that lesion-symptom models trained using adult datasets can be directly translated to predict chronic sensorimotor and language outcomes after childhood stroke.

The relationships captured by adult lesion-symptom models enabled robust outcome predictions in children. When applied to the pediatric lesion data, the adult models generated chronic outcome predictions that correlated with true pediatric impairment ratings (**Figure 3B**), enabled classification of chronic impairment status (**Figure 3C**), and afforded unique explanatory power in multivariable outcome models (**Figure 3D**), with each standard deviation increase in impairment severity predicted by the adult models conferring ∼3-3.5x greater odds of worse chronic outcomes in children when accounting for other factors (**Figure 3D**). Surprisingly, we did not observe a significant advantage for models trained directly in the pediatric cohort (**Figure 4**, **Supplementary Figures 4-5**), and when applied to the same lesion input data, the predictions made by the adult and pediatric models were generally similar to each other (**Figure 5**), suggesting that the adult and pediatric models largely converged on common underlying structure-function relationships despite being trained to predict different outcome measures in developmentally distinct patient samples. These results demonstrate the feasibility of translating adult lesion-symptom models to predict chronic domain-level outcomes after childhood stroke, and suggest that the critical anatomy underlying chronic sensorimotor and language impairments may be generally conserved from early childhood to adulthood.

### Lesion location as a predictor of childhood stroke outcomes

Lesion location is an important predictor of chronic outcomes in adult stroke survivors^12–14^, but findings regarding its value as a predictor of chronic outcomes after childhood stroke have been mixed^9,10^. Anterior circulation infarcts^6,18^, infarcts that involve both small and large vascular territories^3^, infarcts of the lenticulostriate artery^6^, and infarcts affecting both cortical and subcortical structures^6,7^ have been associated with increased risk of poor outcomes after childhood stroke, but findings reported in the literature are inconclusive and lack consistency^3,6,7,10,17,18,46^. This may reflect, at least in part, the limited spatial resolution of lesion measures employed by previous studies on this topic, which have typically relied on very coarse-grained lesion characteristics (e.g. arterial territory, cortical vs. subcortical affection, etc.) rather than on the high-resolution measures of lesion location (i.e. voxel-wise lesion segmentations) that are standard for investigating lesion-symptom relationships in adults^10^. Indeed, we recently conducted an inferential voxel-based lesion-symptom mapping analysis of domain-level outcomes in the same pediatric cohort studied here, and we found robust voxel-level associations with different outcome domains^25^, supporting the view that the coarse-grained summary measures used by prior studies may not be sufficient to enable the identification of robust lesion-symptom associations in children^10^. Here, using both true cross-sample prediction approaches and in-sample cross-validation analyses, we consistently found that lesion location information, whether captured by the adult or pediatric models, provided unique predictive value beyond that attributable to other known risk factors such as lesion size and age at stroke onset, establishing lesion location as an important predictor of chronic childhood stroke outcomes.

### Adult-to-pediatric lesion-symptom model translation

Our primary goal was to evaluate whether the structure-function relationships captured by adult lesion-symptom models can be successfully translated to predict childhood stroke outcomes, motivated in part by our recent work showing that lesion sites associated with post-stroke epilepsy in adults also confer heightened post-stroke epilepsy risk in children^24^. Here, we show that the potential for adult-to-child translation of structure-function relationships is not restricted to pathological outcomes such as epilepsy, but extends to critical domain-level neurological outcomes such as chronic sensorimotor and language impairments. This is an important finding that highlights adult-to-pediatric model translation as a straightforward and accessible approach with the potential to accelerate the development of lesion-informed outcome prediction approaches for childhood stroke and other pediatric conditions. Determining how to further improve and refine this approach is an important next step.

Ensemble modeling or “model stacking” approaches have shown promise for predicting chronic outcomes after adult-onset stroke^12,14,47^. One might therefore envision the development of pediatric ensemble models that combine clinical and demographic variables with lesion-driven predictions obtained from lesion-symptom models trained to predict diverse neurological/neuropsychological outcomes in adults, allowing for comprehensive mapping between adult model-predicted deficit profiles and childhood stroke outcomes. This approach would also allow for the inclusion of pediatric lesion-symptom models as they become available, and could also accommodate models based on lesion-derived connectivity features in addition to lesion location information^13–15,47,48^, providing a flexible and expandable framework for outcome prediction that could be iteratively updated as new datasets, models, and features become available. Alternately, parameters from deep learning models trained using adult datasets could be used to “warm start” analogous pediatric models to improve training efficiency and reduce computational burden, although very large (e.g. N>900) training datasets would be required for such approaches to offer an advantage over traditional machine learning approaches such as those used here^49^. Beyond these examples, our results support the development of novel approaches that leverage adult lesion-symptom relationships to inform pediatric outcome prediction.

### Comparison of adult vs. pediatric lesion-symptom models

Surprisingly, lesion-symptom models trained directly in the pediatric cohort did not significantly outperform those trained using adult data (**Figure 4, Supplementary Figures 4-5**), and when added to the multivariable ordinal logistic regression models, the test fold predictions from the pediatric lesion-symptom models were less strongly associated with chronic impairments (**Supplementary Figure 6**). This supports the validity and robustness of the adult lesion-symptom models, since any biases related to our model training or comparison approaches should favor the pediatric models^27,28^. While the adult models performed well in cross-validation (**Figure 2**), good cross-validation performance does not always confer good generalizability to independent datasets even within the same patient population (e.g. adult-to-adult)^50–54^. Here, the adult models were predicting out-of-sample, across clinical populations, and across outcome measures, while the pediatric models were predicting out-of-fold but within-sample, within-population, and within-outcome. Nonetheless, we observed comparable performance when comparing the pediatric and adult model predictions via cross-validation in the pediatric cohort, suggesting that they independently converged on common underlying anatomical signals present in both children and adults. The observation that the pediatric and adult lesion-symptom models made highly similar predictions when applied to the same lesion inputs further supports this interpretation (**Figure 5**).

It is expected that the effects of injury will differ between developing vs. mature brains^1,2,9^, so why would the adult and pediatric models capture such similar structure-function relationships? One explanation is that the critical anatomical infrastructure underlying sensorimotor and language functions is effectively “in-place” even by early childhood, such that lesion locations producing chronic deficits in a given domain of function are generally conserved across most of the lifespan. This explanation appears largely consistent with recent developmental neuroscience work indicating that a broadly “adult-like” topographic organization of large-scale functional brain networks, including sensorimotor and language networks, is present in young children and even (to a lesser extent) in neonates^21–23^. Similarly, while limited in scale and scope, recent work on neonatal stroke has also identified lesion-symptom relationships that appear to be spatially consistent with those identified in adults^55^.

Importantly, the spatial conservation of critical structures across development would not preclude the possibility that the form and/or severity of impairment following damage to those structures might vary depending on the developmental stage of relevant brain networks at the time of injury^10^. For example, there is evidence suggesting an enhanced capacity of the right hemisphere to take over language functions prior to the consolidation of “dominant” language systems in the left hemisphere^23,56–58^. Along these lines, we recently found that the risk of expressive language deficits conferred by extensive damage to the left hemisphere increases with older age at stroke onset in the same pediatric cohort studied here^25^. However, in the current study, we did not find evidence for a separate moderating effect of age on the relationship between chronic language outcomes and the severity of language impairments predicted by the adult models, suggesting that after accounting for hemisphere-level interactions with onset age, any remaining developmental variation in the effects of lesion location on general language outcomes may be relatively minor, although this should be confirmed by future studies.

Finally, we observed that while, in general, the adult and pediatric lesion-symptom models made highly similar predictions when applied to the same lesion data, they diverged with respect to the overall severity of predicted language outcomes for patients with damage to the right hemisphere (**Figure 5B**). We also observed that when removing shared prediction variance attributable to hemispheric lesion volumes, the correlation between the adult and pediatric model predictions was substantially lower for patients with right unilateral lesions relative to patients with left unilateral lesions, and the deficit-predicting weight topographies of the adult and pediatric models were much more similar in the left hemisphere than in the right hemisphere. While speculative, these observations suggest that the relationship between right hemispheric damage and language outcomes may differ between children vs. adults.

Consistent with this view, right hemispheric lesion volume was a statistically significant predictor in the ordinal logistic regression models of the pediatric language outcome after incorporating the adult lesion-symptom model predictions (**Figure 3D**, **Supplementary Table 1**), but not after incorporating predictions from the pediatric lesion-symptom model (**Supplementary Figure 6**), suggesting that the adult vs. pediatric models differed with respect to how much they captured effects of extensive right hemispheric damage on language outcomes in children. However, the adult model predictions were more strongly associated with pediatric language outcomes (**Figure 3D**, **Supplementary Figure 6**), and the pediatric “expanded” model failed to provide a significant advantage over the adult “expanded” model in cross-validation (**Figure 4B, Supplementary Figure 4**), raising doubts about whether the divergent right hemispheric weight topographies reflect meaningful developmental differences in the effects of right hemispheric damage on chronic language outcomes. It is possible that the observed differences instead reflect other factors such as differences in the sensitivity of the adult vs. pediatric language outcome measures to right hemispheric damage. Thus, while we cannot confidently interpret these observations, they nonetheless provide an example of how model failures or disagreements can reveal potentially interesting insights into potential brain-behavior relationships and support the generation of novel hypotheses that can be refined and tested in subsequent work^59^.

### Limitations

This study has several limitations. First, our pediatric sample did not include neonatal/perinatal stroke, and so our conclusions only apply to childhood stroke. Second, our pediatric outcome measures were derived from the sensorimotor and language domain scores of the PSOM/RRQ and therefore lack granularity. Even so, PSOM scores have been shown to exhibit robust associations with quantitative measures of neuropsychological test performance^33^, and we found that adult models trained using more granular neuropsychological outcomes were able to achieve excellent cross-outcome and cross-cohort generalization to outcome measures derived from the PSOM/RRQ. Time intervals between stroke onset and neurological assessment also varied in our pediatric cohort, and so some variability in outcomes could be attributable to variation in the interval between onset and assessment, although we attempted to mitigate this by including it as a covariate in our analyses.

### Conclusions

Structure-function relationships captured by lesion-symptom models trained in patients with adult-onset brain lesions can be translated to predict chronic sensorimotor and language outcomes in childhood stroke survivors. Lesion-symptom models trained on data from adults vs. children largely converge on common underlying anatomical signals, suggesting a general conservation of critical structures from childhood to adulthood. Adult-to-pediatric model translation provides a potential means of accelerating lesion-informed outcome prediction in pediatric populations.

## Data Availability

Data are available on request to the senior co-authors or via submitting an International Pediatric Stroke Study (IPSS) research proposal form, available at https://internationalpediatricstroke.org/ipss-research/.

Analysis scripts, along with MATLAB Live notebooks showing how each result in the main text and supplement was generated, are available at https://github.com/jcgriffis/adult_to_pediatric_lsm_translation. Trained adult and pediatric lesion-symptom models will be made available upon publication.

## Data Availability

Data are available on request to the senior co-authors or via submitting an International Pediatric Stroke Study (IPSS) research proposal form, available at https://internationalpediatricstroke.org/ipss-research/.
Analysis scripts, along with MATLAB Live notebooks showing how each result in the main text and supplement was generated, are available at https://github.com/jcgriffis/adult_to_pediatric_lsm_translation. Trained adult and pediatric lesion-symptom models will be made available upon publication.

https://github.com/jcgriffis/adult_to_pediatric_lsm_translation

## Acknowledgments

We would like to thank Maya Dhar as the SCOUTS senior research coordinator, the research coordinators at all SCOUTS sites, and the patients and their families for participation in the VIPS and SCOUTS studies.

## Appendix

**Coinvestigators**

| Name | Location | Role | Contribution |
|---|---|---|---|
| Clara Di Germiano | Vitalant Research Institute, San Francisco, CA | Co-Investigator | Laboratory Core |
| Philip J. Norris | Vitalant Research Institute, San Francisco, CA | Co-Investigator | Laboratory Core Lead |
| Adam L. Numis | UCSF, San Francisco, CA | Co-Investigator | Epileptologist |
| Lauren A. Beslow | Children’s Hospital of Philadelphia, Philadelphia, PA | Site Investigator | Enrollment and data collection |
| Paola Pergami | Children’s National Hospital, Washington DC | Site Investigator | Enrollment and data collection |
| Rachelle Herring | Cook Children’s Hospital, Fort Worth, TX | Site Investigator | Enrollment and data collection |
| Jenny Wilson | Oregon Health & Science University, Portland, OR | Site Investigator | Enrollment and data collection |
| Catherine Amlie-Lefond | Seattle Children’s Hospital, Seattle, WA | Site Investigator | Enrollment and data collection |
| Neil Friedman | Phoenix Children’s Hospital, Phoenix, AZ | Site Investigator | Enrollment and data collection |
| Marcie Files | Children’s Mercy Hospital, Kansas City, MO | Site Investigator | Enrollment and data collection |
| Beth Anne Cavanaugh | Le Bonheur Children’s Hospital, Memphis, TN | Site Investigator | Enrollment and data collection |
| Ryan Felling | Johns Hopkins Children’s Center, Baltimore, MD | Site Investigator | Enrollment and data collection |
| Lisa Sun | Johns Hopkins Children’s Center, Baltimore, MD | Site Investigator | Enrollment and data collection |
| Andrea Pardo | Lurie Children’s Hospital of Chicago, Chicago, IL | Site Investigator | Enrollment and data collection |

