## Supplementary Material for "Predicting pediatric stroke outcomes using adult lesion-symptom models"

**Supplementary Table 1.** Multivariable Ordinal Logistic Regression Models

| SM “Base” Model | | | | | |
| --- | --- | --- | --- | --- | --- |
| Predictor | **Std. Beta** | **aOR** | **95% CI** | **t-statistic** | ***P*-value** |
| LH Volume | 0.59 | 1.80 | 1.50, 2.16 | 6.28 | 3.33e-10 |
| RH Volume | 0.45 | 1.59 | 1.30, 1.94 | 4.57 | 4.83e-06 |
| Age at Stroke | -0.13 | 0.88 | 0.73, 1.04 | -1.48 | 0.14 |
| Time to Test | 0.07 | 1.07 | 0.90, 1.28 | 0.81 | 0.42 |
| SM “Expanded” Model | | | | | |
| Predictor | **Std. Beta** | **aOR** | **95% CI** | **t-statistic** | ***P*-value** |
| LH Volume | -0.14 | 0.87 | 0.66, 1.16 | -0.94 | 0.35 |
| RH Volume | -0.37 | 0.69 | 0.51, 0.93 | -2.40 | 0.02 |
| Age at Stroke | -0.21 | 0.81 | 0.67, 0.97 | -2.30 | 0.02 |
| Time to Test | 0.09 | 1.10 | 0.92, 1.31 | 1.04 | 0.30 |
| Adult Model | 1.27 | 3.55 | 2.43, 5.19 | 6.57 | 4.98e-11 |
| LANG “Base” Model | | | | | |
| Predictor | **Std. Beta** | **aOR** | **95% CI** | **t-statistic** | ***P*-value** |
| LH Volume | 0.65 | 1.92 | 1.59, 2.32 | 6.76 | 1.41e-11 |
| RH Volume | 0.33 | 1.39 | 1.15, 1.68 | 3.46 | 0.0005 |
| Age at Stroke | -0.25 | 0.78 | 0.63, 0.97 | -2.22 | 0.03 |
| Time to Test | 0.17 | 1.19 | 0.97, 1.45 | 1.64 | 0.10 |
| LANG “Expanded” Model | | | | | |
| Predictor | **Std. Beta** | **aOR** | **95% CI** | **t-statistic** | ***P*-value** |
| LH Volume | -0.41 | 0.67 | 0.37, 1.19 | -1.38 | 0.17 |
| RH Volume | 0.30 | 1.35 | 1.13, 1.61 | 3.29 | 0.001 |
| Age at Stroke | -0.30 | 0.74 | 0.59, 0.93 | -2.61 | 0.009 |
| Time to Test | 0.15 | 1.17 | 0.95, 1.43 | 1.48 | 0.14 |
| Adult Model | 1.10 | 3.0 | 1.67, 5.30 | 3.70 | 0.0002 |

Std. Beta – standardized beta coefficient, aOR - adjusted cumulative odds ratio, CI – confidence interval

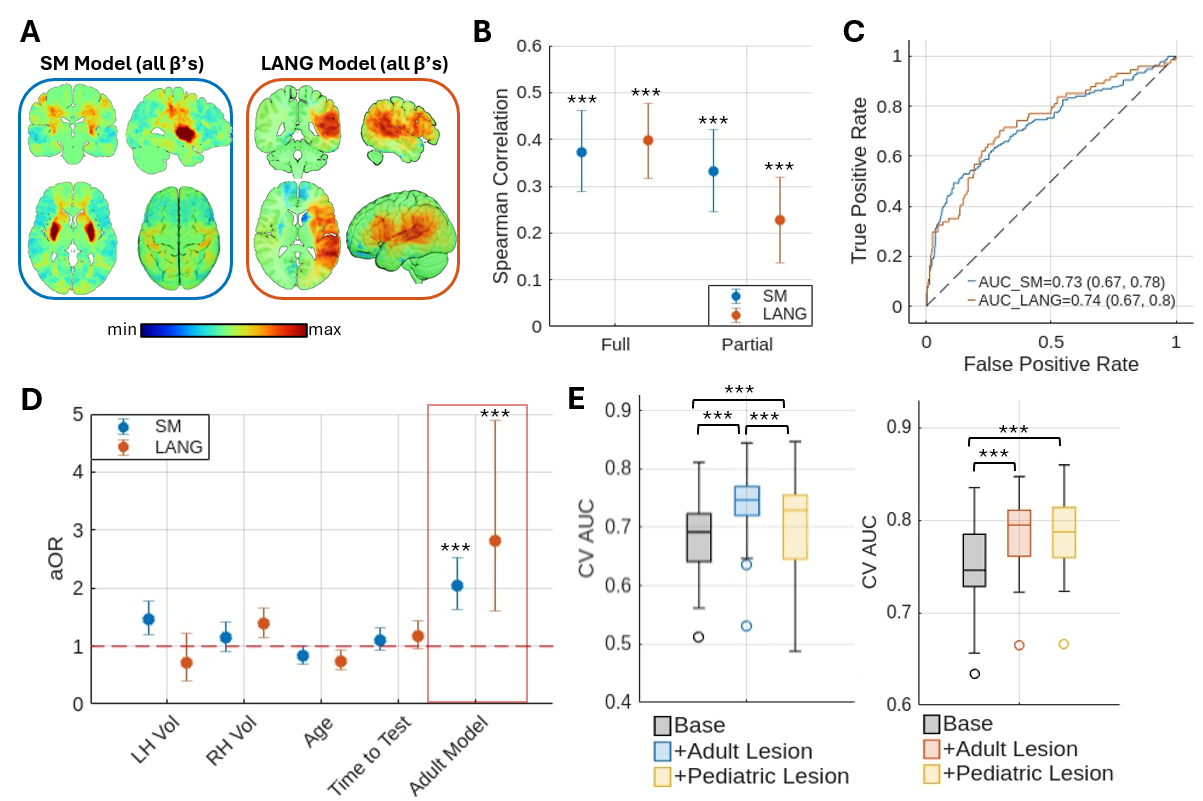

**Supplementary Figure 1.** Analyses using full (i.e. positive and negative) model weights to generate outcome predictions. **A.** Full model weights (i.e. beta coefficients) are shown for the sensorimotor (SM) and language (LANG) models. Slices are shown in radiological convention (i.e. left is on the right). **B**. Full and partial Spearman rank correlations between the adult-derived model predictions of sensorimotor (blue – SM) and language (orange – LANG) outcomes and true observed outcomes in the pediatric stroke dataset are shown along with corresponding 95% CIs. **C.** Receiver-operating characteristic (ROC) curves for discriminating between patients with vs. without sensorimotor (blue) and language (orange) impairment constructed using the adult-derived model predictions generated for the pediatric stroke dataset are shown. Areas under the curve (AUCs) and 95% confidence intervals (CIs) are shown in the legend. **D**. Adjusted odds ratios (aOR) for standardized predictors from multivariable ordinal logistic regression models of each outcome (blue – SM, orange – LANG) that included the adult-derived model predictions (red box) are shown along with 95% CIs. **E.** Boxplots show comparisons of cross-validation classification performance estimates (AUC) between the “base” logistic ridge regression models (grey), “expanded” models that included the adult-derived model predictions (blue – SM, orange – LANG), and “expanded” models that included test-fold predictions obtained from cross-validation of pediatric lesion-symptom models (yellow). Higher values indicate better test fold classification performance. All statistically significant effects from the main analysis remained statistically significant when using the full model weights to generate predictions, although some effects were weaker when using the full model predictions. ****P_FWE_*<0.001

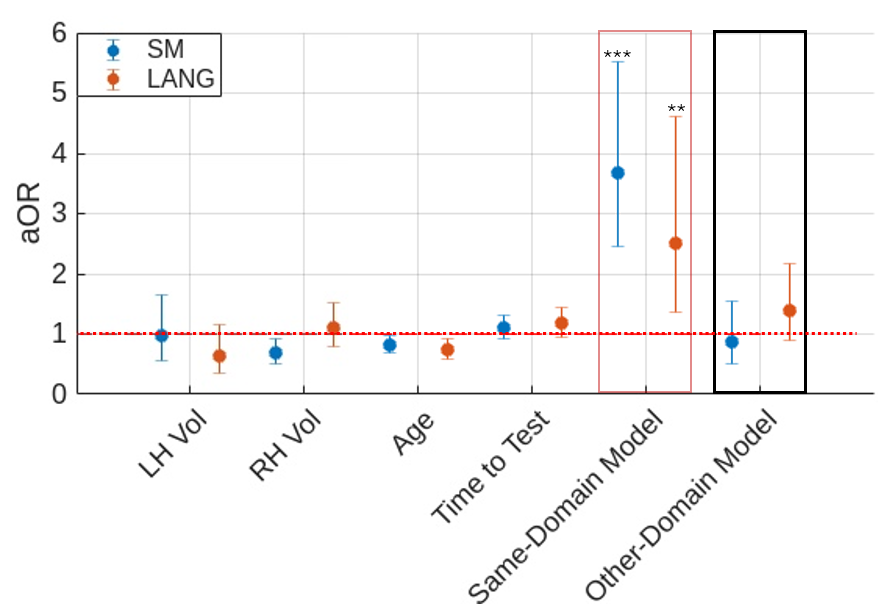

**Supplementary Figure 2.** Domain specificity of adult-to-pediatric lesion-symptom model translation. For each outcome, predictions from the domain-mismatched adult lesion-symptom model (i.e. Token Test model for SM, Grooved Peg-Board model for LANG) were added to the “expanded” model that included the domain-matched (i.e. Grooved Peg-Board model for SM, Token Test model for LANG) adult model predictions. Standardized aORs and 95% CIs are shown for the predictors in each model. The domain-matched adult lesion-symptom model predictions (i.e. Same-Domain Model) remained significant predictors of each outcome after adding the domain-mismatched predictions, but adding the domain-mismatched adult lesion-symptom model predictions (i.e. Other-Domain Model) did not significantly improve model fits for either outcome (SM: Chi-square[1]=0.20, *P*=0.67; LANG: Chi-square[1]=2.21, *P*=0.14), and the domain-mismatched predictions were not significant in either model. ****P*<0.001, ***P*<0.005

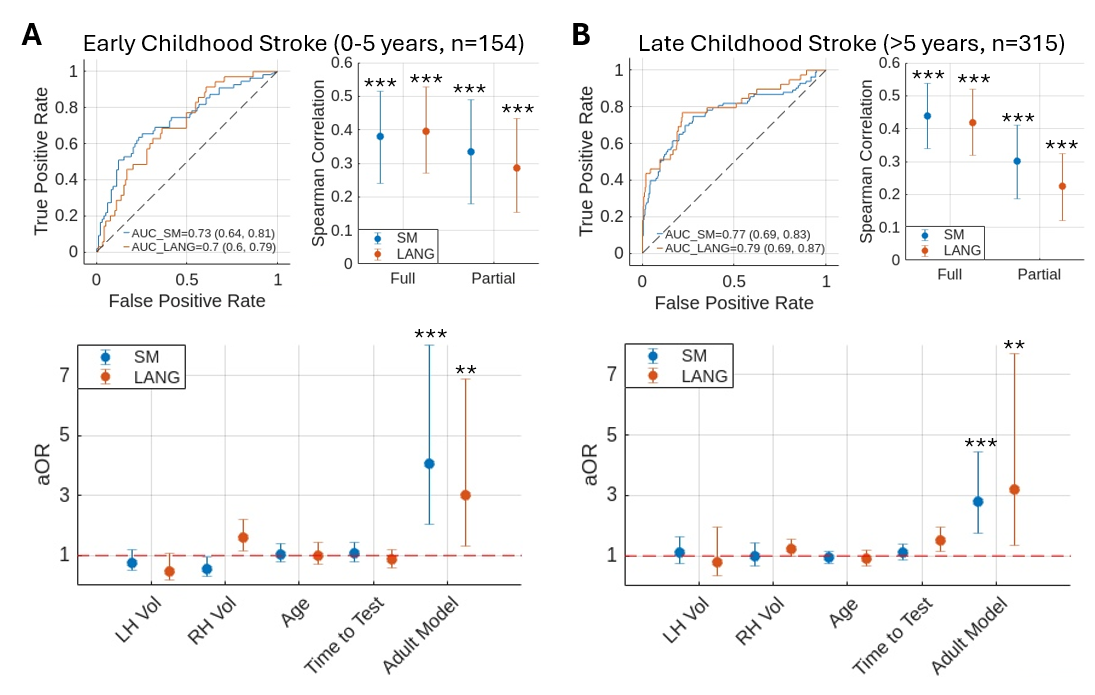

**Supplementary Figure 3.** Translating adult lesion-symptom models to separately predict outcomes after early and late childhood stroke. The primary adult-to-pediatric outcome prediction analyses from Figure 3 of the main text were repeated separately for **(A)** early childhood stroke survivors (i.e. age at stroke onset between 0-5 years, n=154) and **(B)** late childhood stroke survivors (i.e. age at stroke onset > 5 years, n=315). All statistically significant results obtained in the analyses of the full pediatric cohort were also obtained in the separate analyses of early vs. late childhood stroke survivors. ****P* <0.001, ***P* <0.01

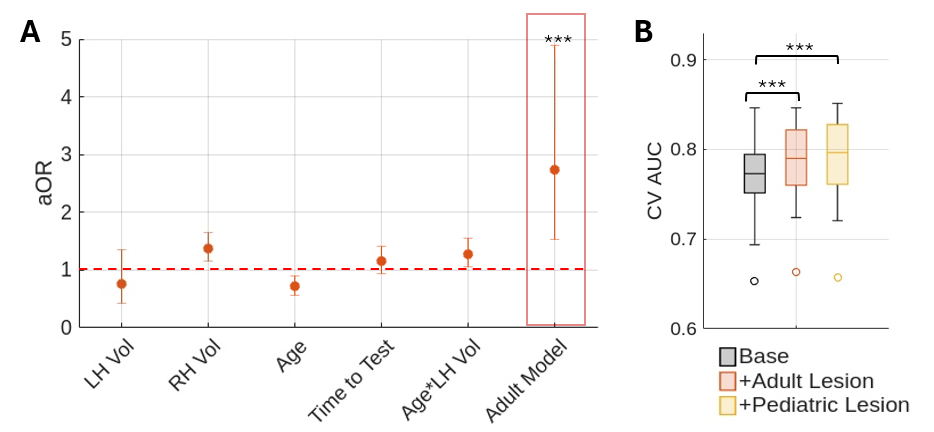

**Supplementary Figure 4.** Language model results when including an interaction term between age at stroke onset and left hemispheric lesion volume. **A.** Standardized aORs are shown for each predictor in the multivariable ordinal logistic regression model of the pediatric language outcome that includes the interaction term between age and left hemispheric lesion volume. When the interaction term was included in the model, adding the adult model predictions significantly improved the model fit (Chi-square[1]=12.79, *P*=0.0003), and the effect of the adult model predictions remained statistically significant. **B**. Boxplots show cross-validation AUCs (CV AUC) for the logistic ridge regression models predicting chronic pediatric language impairment status. All models are the same as for the LANG models reported in Figures 3 and 4 of the main text, but additionally include the interaction term between age at stroke onset and left hemispheric lesion volume. Adding the adult lesion-symptom model predictions to the base model that includes the interaction term still significantly improves the cross-validation performance (t[24]=5.62, *P*=8.65e-06), as does adding the pediatric lesion-symptom model predictions (t[24]=6.07, *P*=2.86e-06), and differences between the expanded models including the adult vs. pediatric lesion-symptom model predictions remain non-significant (t[24]=-1.77, *P*=0.09).

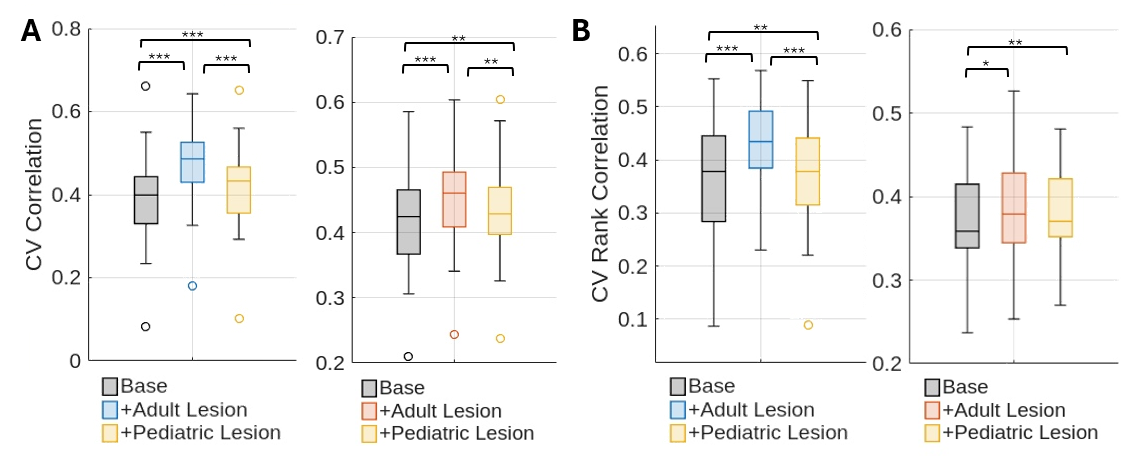

**Supplementary Figure 5.** Cross-validation comparisons using ridge regression models. The cross-validation model comparison analysis was performed using ridge regressions to predict the pediatric impairment scores instead of classifying pediatric impairment status. Results are shown for the sensorimotor (left within each panel) and language (right within each panel) outcomes. Model performance was measured in terms of the correlation between predicted and observed impairment scores for each test fold. Box plots show distributions of cross-validation correlations for all test folds. “Expanded” models that included the adult lesion-symptom model predictions significantly outperformed both the “base” models and, for several analyses, the alternate “expanded” models that included the pediatric lesion-symptom model predictions. **A.** Results are shown using Pearson correlation to summarize CV performance. **B.** Results are shown using Spearman correlation to summarize performance. ****P* <0.001, ***P*<0.01, **P*<0.05

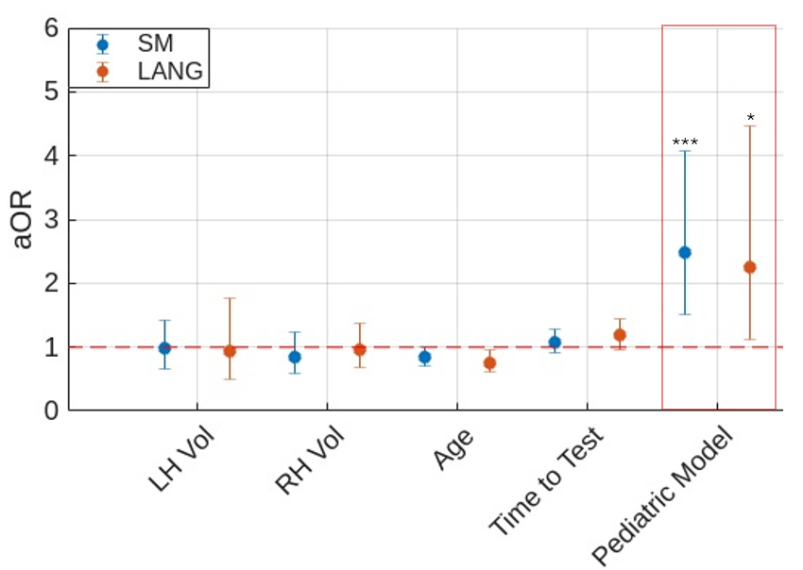

**Supplementary Figure 6.** Effects of adding pediatric lesion-symptom model test fold predictions to the “base” ordinal logistic regression model. The average (across repetitions) test fold prediction for each pediatric patient was added to the “base” multivariable ordinal logistic regression model (i.e. instead of the adult model predictions). Adding the standardized pediatric lesion-symptom model predictions significantly improved both the SM (Chi-square[1]=12.75, *P*=3.56e-4) and LANG (Chi-square[1]=5.42, *P*=0.02) models, and the pediatric model predictions were significant predictors in each model, although aORs for the pediatric model predictions were weaker than those for the adult model predictions (i.e. Figure 3D). *** *P*<0.001, * *P*<0.05
